# Co-expression in tissue-specific gene networks links genes in cancer-susceptibility loci to known somatic driver genes

**DOI:** 10.1101/2023.09.08.23295254

**Authors:** Carlos G. Urzúa-Traslaviña, Tijs van Lieshout, Floranne Boulogne, Kevin Domanegg, Mahmoud Zidan, Olivier B. Bakker, Annique Claringbould, Jeroen de Ridder, Wilbert Zwart, Harm-Jan Westra, Patrick Deelen, Lude Franke

## Abstract

**Background:** The genetic background of cancer remains complex and challenging to integrate. Many somatic mutations in genes are known to cause and drive cancer, while genome-wide association studies (GWAS) of cancer have revealed many germline risk factors associated with cancer. However, the overlap between known somatic driver genes and positional candidate genes from GWAS loci is surprisingly small. We hypothesised that genes from multiple independent cancer GWAS loci should show tissue-specific co-regulation patterns that converge on cancer-specific driver genes.

**Results:** We studied recent well powered GWAS of breast, prostate, colorectal and skin cancer by estimating co-expression between genes and subsequently prioritising genes that show co-expression with genes mapping within susceptibility loci from cancer GWAS. We observed that the prioritised genes were strongly enriched for cancer drivers defined by COSMIC, intOGen and Dietlein *et al*. The enrichment of known cancer driver genes was most significant when using co-expression networks derived from non-cancer samples from the relevant tissue of origin.

**Conclusion:** We show how genes in risk loci identified by cancer GWAS can be linked to known cancer driver genes through tissue-specific co-expression networks. This provides an important explanation for why seemingly unrelated sets of genes that harbour either germline risk factors or somatic mutations can eventually cause the same type of disease.

## Background

Cancer continues to be a formidable health challenge, responsible for a substantial burden of morbidity and mortality worldwide (1). Despite the many advances in cancer research, the molecular mechanisms that underlie cancer initiation and progression remain incompletely understood. It is now widely recognised that genetic alterations play a central role in the aetiology of some cancer types, enabling the acquisition of hallmark characteristics such as uncontrolled cell proliferation, evasion of cell death and the ability to invade and metastasise (2). Two distinct types of genetic alterations have emerged as key players in cancer development: germline risk variants and somatic mutations. Germline risk variants are constitutional alterations present in every cell of an individual’s body that are inherited from one or both parents. These mutations can confer an increased susceptibility to developing cancer (3,4). In contrast, somatic mutations arise in somatic cells and progressively accumulate over time, leading to the transformation of normal cells into their malignant counterparts. These kinds of mutations are the driving force behind the clonal expansion of malignant cells and the heterogeneity observed within tumours (5).

These two types of genetic alterations sometimes occur in the same gene suggesting a common mechanism for initiation (cancer susceptibility gene) and sustaining of cancer (cancer driver gene) (6). Here we refer to ‘cancer driver gene’ as genes annotated to drive cancer by the COSMIC CGC, or with evidence of non-neutral somatic mutation frequencies by intOGen and Dietlein *et al* (5,7,8). For example, *TP53* is often somatically mutated in several forms of cancer, but it also harbours rare germline variants that increase the risk of cancer development in Li-Fraumeni Syndrome patients (9). Similarly, studies of rare germline coding variants in breast cancer have also found overlap with the cancer-driver genes derived from tumour data (10).

However, little overlap has been observed between cancer driver genes and genes that are influenced by more common germline variants. Hundreds of susceptibility variants, each conferring a small risk, have now been identified through genome-wide association studies (GWAS) of various cancers (11). Yet only 5% of the genes implicated through cancer GWAS are established cancer driver genes (12).

These observations suggest that common germline cancer risk variants confer their risk by preferentially acting in a set of ‘co-driver’ genes that indirectly facilitate oncogenesis later in life (13). In support of this idea recent studies have found associations between germline variants and the tumour genome of patients with cancer (14,15). However, the question remains if and how the genes affected by common variants are influencing the activity of cancer drivers. As observed with rare germline variants, one possibility is that common germline variants influence the type of somatic mutational pattern of the tumour which then favors the mutation of different cancer drivers (16). Another possibility is that the genes affected by common variants are ‘peripheral’ genes which regulate a set of ‘core’ genes that mediate the cancer risk as suggested by the omnigenic model (17). In line with this model, we hypothesised that some somatic cancer drivers may be regulated by the peripheral genes derived from cancer risk loci identified through GWAS.

We therefore aimed to test this hypothesis for four types of cancer—breast cancer, prostate cancer, colorectal cancer and skin cancer—for which GWAS have been conducted in a substantial number of cases. To achieve this aim, we first ascertained whether the genes inside these GWAS loci are suspected to be functionally related within a gene-network derived from mRNA co-expression. We then determined if ‘core’ genes, genes that are highly linked to genes inside these cancer GWAS loci, are known somatic cancer driver genes. For this purpose, we applied Downstreamer: a gene prioritisation methodology that uses co-expression information to prioritise genes that are significantly co-expressed with genes in significant GWAS loci (18). We explored different gene co-expression networks to determine which regulatory context produces the strongest enrichment of somatic cancer driver genes. We considered co-expression networks generated from a mix of tissues (a multi-tissue network) and from specific tissues. Because cancer and adjacent healthy tissues may have different mRNA expression characteristics (19), we also studied whether networks derived exclusively from cancer or non-cancer samples affect the enrichment of genes with known somatic mutations.

We observed that our approach produces the best results in terms of enrichments for cancer driver genes when using networks derived from non-cancer tissue the same origin as the cancer in question. Overall, our results suggest that genes in cancer GWAS loci seem to converge on downstream cancer driver genes for some cancers, suggesting that genes in GWAS loci may confer their risk by indirectly acting on those genes. Moreover, the prioritisation performed by Downstreamer points to other genes that could be considered novel cancer susceptibility genes.

## Results

We collected summary statistics for 109 different GWAS of cancer traits and focused on those studies that identified at least 90 risk loci and had the highest number of cases among all the GWAS of the same cancer type (Table S1). After selection, four cancer GWAS remained that studied the following cancers: prostate, breast, colon and skin (Table 1).

**Table 1:** Overview of the four genome-wide association studies (GWAS) used in this study. GWAS ID is the identifier in the GWAS catalogue. The number of independent loci was determined using the Downstreamer methodology (see Methods). Number of cases is the number of European ancestry cases reported by the GWAS catalogue.

| Study | GWAS ID | Cancer | Number of independent loci | Number of cases |
| --- | --- | --- | --- | --- |
| Schumacher *et al.*, *Nat. Genet.* 2018 | GCST006085 | Prostate | 335 | 79,148 |
| Zhang *et al.*, *Nat. Genet.* 2020 | GCST010098 | Breast | 348 | 133,384 |
| Sakaue *et al.*, *Nat. Genet.* 2021 | GCST90018921 | Skin | 123 | 25,928 |
| Fernandez-Rozadilla *et al.*, *Nat. Genet.* 2022 | GCST90102485 | Colorectal | 99 | 100,204 |

We next created a catalogue of confirmed and putative somatic cancer driver genes for a specific tissue of origin by combining genes annotated to drive cancer by the COSMIC CGC (Tier 1 and Tier2), or with evidence of non-neutral somatic mutation frequencies by intOGen and Dietlein *et al* (5,7,8) (Fig. S1, Table 2, Table S2). Using these resources, we evaluated if cancer drivers are highly coregulated with genes inside GWAS loci (Figure 1).

**Figure 1:**
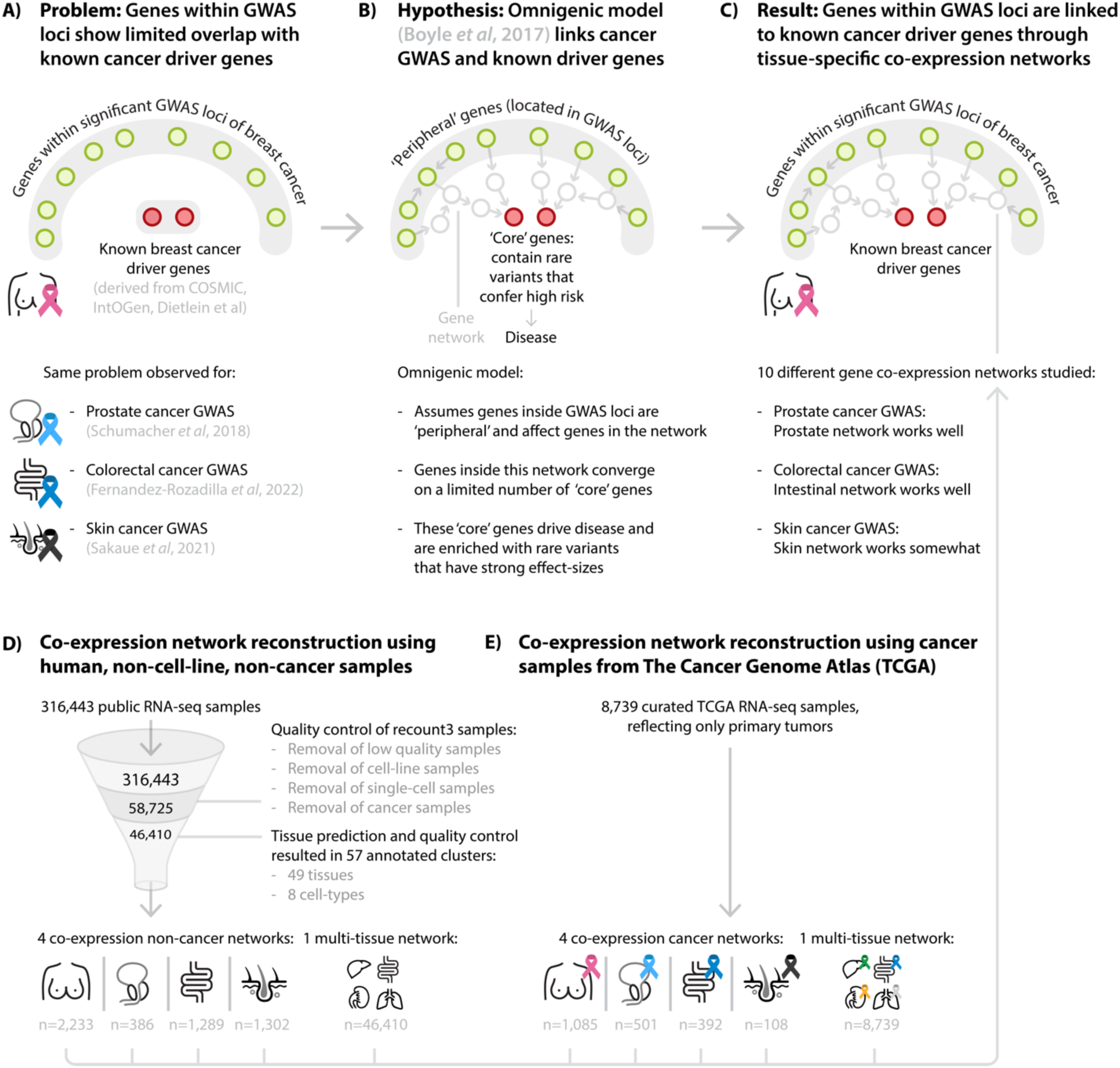
Overview of application of gene-prioritisation methods on cancer GWAS summary statistics to identify cancer-type-specific somatic driver genes.

**Table 2:** Cancer driver genes per GWAS.

| Tissue of origin | Total | Unique to tissue |
| --- | --- | --- |
| Breast | 131 | 51 |
| Prostate | 106 | 49 |
| Colon | 142 | 71 |
| Skin | 177 | 94 |

### Cancer-specific driver genes show limited enrichment for cancer GWAS signal

Previous reports have indicated that there is limited overlap between somatic cancer drivers and genes in cancer GWAS loci (12). Because we are studying very recent GWAS that could have identified additional genes since the previous report, we first re-evaluated this overlap. To do so, we used PascalX (20) to estimate gene-level p-values from the GWAS summary statistics of the prostate, breast, skin and colon cancer studies using 19,922 protein-coding genes and a window size of 25kb around each gene (Table S3). The resulting gene-level p-values denote to what extent variants within and near these genes show an association signal in the GWAS. To test for the expected baseline enrichment, we looked at how many of the Bonferroni significant (p-value < 2.51 × 10^-6^) PascalX prioritized genes are known tissue-specific somatic cancer drivers. Among the Bonferroni significant hits, 9 out of the 344 (2.62%) genes prioritised for the breast cancer GWAS were known somatic tissue-specific cancer drivers (*FGFR2*, *MAP3K1*, *RAD51B, ESR1, CDKN2A, CASP8, TBX3, AKT1* and *PIK3R1,* Fisher exact p-value: 0.00041, median distance to nearest index variant = 34.9 Kb). For the prostate cancer GWAS 5 out of the 382 (1.31%) genes prioritized by PascalX are known prostate cancer drivers (*KLK2, KLK3, SPEN, TMPRSS2* and *RNF43,* Fisher exact p-value: 0.05, median distance to nearest index variant = 6.2 Kb). In the PascalX prioritization for the skin cancer GWAS 4 out of the 148 (2.70%) genes prioritised are known skin cancer drivers (*CASP8, TERT, CDKN2A* and *BCL2L12,* Fisher exact p-value: 0.04, median distance to nearest index variant = 40.8 Kb). No Bonferroni significant cancer drivers were found for the colorectal cancer GWAS in the PascalX analysis. We also tested (one-sided Wilcoxon signed-rank test) whether all cancer-specific driver genes generally show a more significant gene-level GWAS p-value compared to all other genes (Fig. S2). We observed significant enrichments for the breast (p-value: 1.13 × 10^-5^) and colon cancer GWAS (p-value: 0.008), suggesting that beyond the Bonferroni significant hits there is further enrichment for the breast cancer and colon cancer GWAS. These findings suggest that, in general, PascalX identifies some cancer driver genes using summary statistics of well-powered GWAS and that the amount of cancer drivers is consistent with the percentages reported in the literature (12). However, many more somatic drivers associated with these cancers are not called as significant by PascalX.

### Additional cancer driver genes can be linked to GWAS loci through co-expression networks

Following the low overlap between genes identified by PascalX from GWAS and somatic driver genes we next tested the hypothesis of whether genes identified by PascalX influence cancer drivers indirectly. For this, we employed Downstreamer, which uses a gene network (co-expression model derived from bulk mRNA transcription) together with gene level p-values derived by PascalX. Downstreamer then calculates a key gene prioritisation score that captures if a gene is significantly co-expressed with genes prioritized by PascalX from GWAS data (18).

Co-expression networks derived from a mixture of tissues are derived from a comparatively larger number of samples and therefore may better model co-expression links between genes due to better statistical power. However, deriving networks from tissue-specific subsets of samples may result in networks that better capture co-expression links relevant to the tissue. We therefore tested networks derived exclusively from the tissue of origin of the primary tumour or from a mixture of multiple tissues using publicly available data from the cancer genome atlas (TCGA) and recount3 (21) (Table 3). Each co-expression network has a different number of components that capture the variation in transcriptional patterns which Downstreamer uses when performing the prioritizations (18). Additionally, as cancer predisposition mechanisms may operate both before and after the onset of oncogenesis, we compared the results from networks derived from cancer tissue and non-cancer tissue. For every option of multi-tissue and tissue-specific networks, we applied the Downstreamer methodology to the cancer GWAS (Table S4).

**Table 3:** Co-regulation networks generated in this study.

| Co-regulation network | Resource | Components | Genes | Samples | Type |
| --- | --- | --- | --- | --- | --- |
| Non-cancer adipose breast | recount3 | 223 | 33,935 | 2,233 | Tissue-specific non-cancer |
| Non-cancer colon | recount3 | 139 | 35,623 | 1,289 | Tissue-specific non-cancer |
| Non-cancer prostate | recount3 | 63 | 33,873 | 386 | Tissue-specific non-cancer |
| Non-cancer skin | recount3 | 141 | 35,524 | 1,302 | Tissue-specific non-cancer |
| Breast cancer | TCGA | 161 | 35,523 | 1,085 | Tissue-specific cancer |
| Colorectal cancer | TCGA | 65 | 35,523 | 392 | Tissue-specific cancer |
| Prostate cancer | TCGA | 81 | 35,523 | 501 | Tissue-specific cancer |
| Skin cancer | TCGA | 27 | 35,523 | 108 | Tissue-specific cancer |
| Multi-tissue non-cancer | recount3 | 848 | 28,942 | 46,410 | Non-cancer multi-tissue |
| Multi-tissue cancer | TCGA | 1,864 | 35,523 | 8,739 | Cancer multi-tissue |

We first describe the results of applying Downstreamer using the two multi-tissue networks: one derived exclusively from cancer (TCGA multi-cancer) and one derived from non-cancer tissue data (recount3 non-cancer multi-tissue; Table 3, Fig. S3).

Among the prioritised genes we identified known somatic driver genes passing the Bonferroni significance threshold (p < 0.05/19,922 = 2.51x10^-6^). The result from the analysis with the cancer multi-tissue network and the prostate cancer GWAS analysis included the gene *CANT1* (p-value: 2.0 × 10^-6^). *CANT1* is located 7.7Mb from the nearest prostate cancer GWAS index variant. The large distance from the nearest index variant suggests that the GWAS signal near this gene is weak and explains why this gene was not genome wide significant through the PascalX approach (PascalX p-value: 0.043). *CANT1*encodes the androgen-regulated protein *CANT1*, which has previously been found to form a fusion transcript with *ETV4* in prostate cancer (22,23). Note that *ETV4* does not seem to be prioritized by PascalX (PascalX p-value: 0.54) or Downstreamer (p- value 0.64). Results from the analysis with the non-cancer multi-tissue network and the colon cancer GWAS included the genes *KMT2B, PTBP1* and *GEN1* (p-values 2.0 × 10^-6^, 3.0 × 10^-6^ and 3.0 × 10^-6^, respectively). Each of these genes are located at least 2.5Mb away from the nearest index variant. *PTBP1* expression has previously been associated with invasiveness in colorectal cancer through alternative splicing of cortactin (24). The analysis of the breast cancer GWAS with the same non-cancer multi-tissue network resulted in one Bonferroni significant gene *LRP1* (p-values 1.22 × 10^-6^). *LRP1* repressed xenografts of triple negative breast cancer cell lines have been shown to decrease tumour growth and angiogenesis (25). No Bonferroni significant hits were found for the skin cancer GWAS using both cancer and non-cancer multi-tissue networks. To determine if there is overrepresentation of cancer driver genes in the Downstreamer prioritisation scores, we performed a Wilcoxon signed-rank test of cancer specific driver genes versus all other genes. We observed improved enrichment p-values for known somatic cancer drivers in the gene prioritisations for the prostate GWAS when using the cancer multi-tissue network compared with the equivalent enrichment performed on PascalX results of the same GWAS (p-value: 7.76 × 10^-4^ vs. 0.08 one-sided Wilcoxon signed-rank test). We observed a smaller improvement in enrichment for the colon cancer GWAS when using the non-cancer multi-tissue network as compared to PascalX (p-value: 8.43 × 10^-4^ vs. 0.009).

We then created tissue-specific networks derived from cancer and non-cancer samples for prostate, breast, skin and colon tissues. In contrast to the previous multi-tissue networks, these are derived from a smaller subset of tissue specific samples to better capture tissue specific co-expression. We applied Downstreamer to each cancer GWAS using the tissue-specific network corresponding to the tissue of origin (referred to hereafter as a ‘matched network’) (Figure 2, Fig. S4). This analysis also resulted in Bonferroni significant hits: In the matched non-cancer tissue-specific network analysis, we prioritised three genes for breast cancer, *PHLPP1, PUM1* and *CCL28* (p-values: 5.93 × 10^-7^, 1.55 × 10^-6^ and 2.01 × 10^-6^, respectively), and two for colon cancer, *ZZEF1* and *BAZ2A* (p-values: 1.94 × 10^-6^ and 2.56 × 10^-6^, respectively). For breast cancer, there is evidence that *CCL28* and *PHLPP1* may be drivers of breast cancer through the MAPK and AKT pathways, respectively (26,27). While *PUM1* and *BAZ2A* have, to our knowledge, not been implicated for the matching cancer types, they have been shown to be potential drivers in other cancers (28,29).

**Figure 2:**
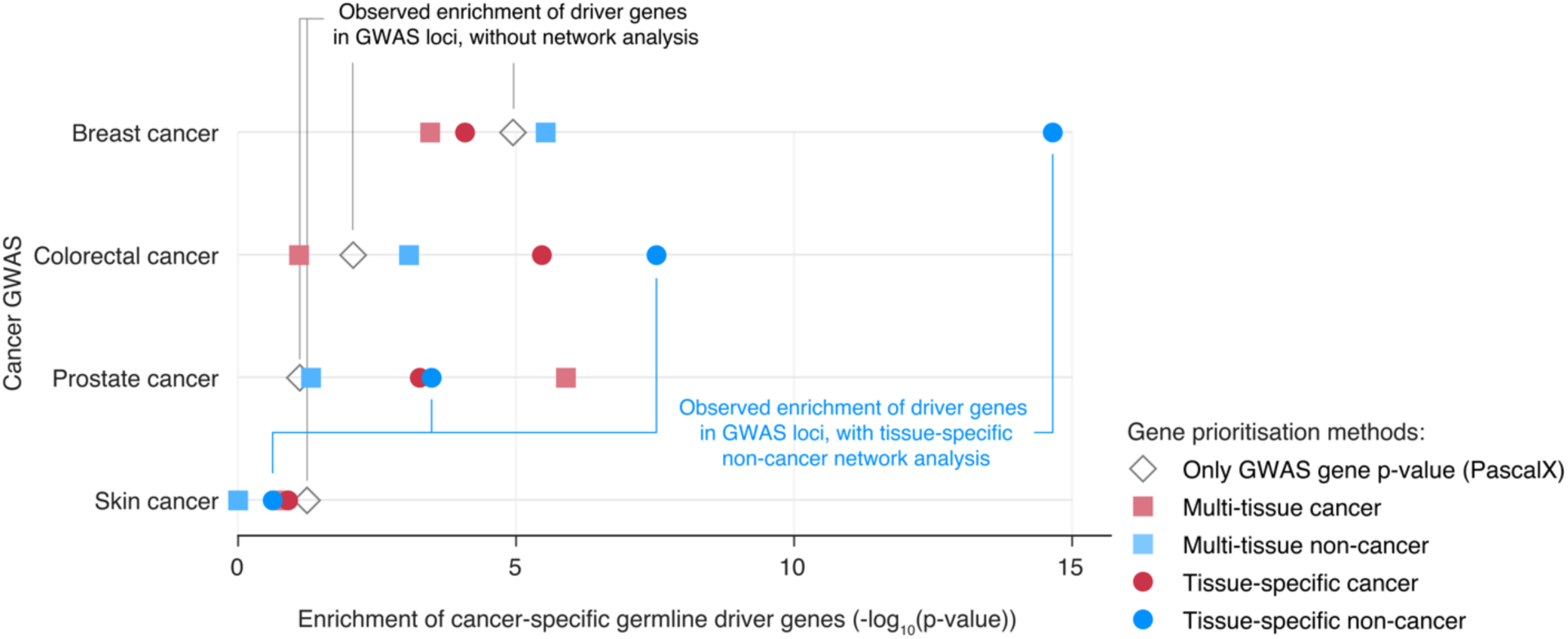
Enrichment of cancer-specific somatic driver genes for different multi-tissue and tissue-specific gene-prioritisation methods applied to cancer GWAS summary statistics. A one-sided Wilcoxon signed-rank test was used to calculate the enrichment of cancer-specific driver genes in the list of prioritised genes (x-axis) for four cancer GWAS studies (y-axis). Open diamonds indicate gene enrichment values for the original GWAS, calculated with PascalX from the GWAS summary statistics . Coloured symbols indicate the enrichments for the different tissue networks, calculated using Downstreamer.

Our selection of driver genes contains both confirmed and putative cancer drivers that have not been validated, we therefore repeated the enrichment analysis while restricting the genes to the high confidence set defined as Tier 1 in the COSMIC Cancer Gene Census. We observed a less significant but comparable enrichment of Tier 1 breast cancer drivers giving support to the results observed in the full set of cancer drivers (Fig. S5). Overall, these enrichments further suggest how GWAS-associated genes may be converging to cancer-associated genes downstream within tissue-specific co-expression networks.

When again performing Wilcoxon ranked sum test to test the enrichment of cancer specific driver genes versus other genes, we observed that the breast and colon cancer GWAS showed stronger enrichments using the matched tissue-specific networks compared to multi-tissue networks (see p-values in Table 4).

**Table 4.**
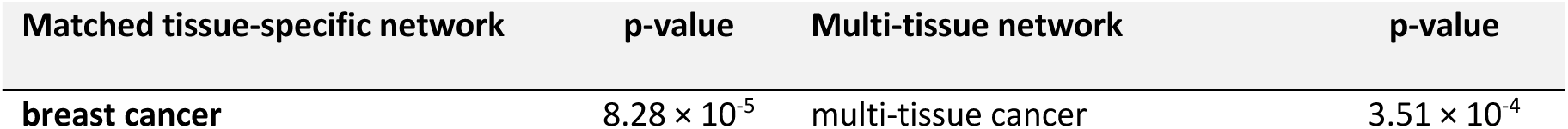
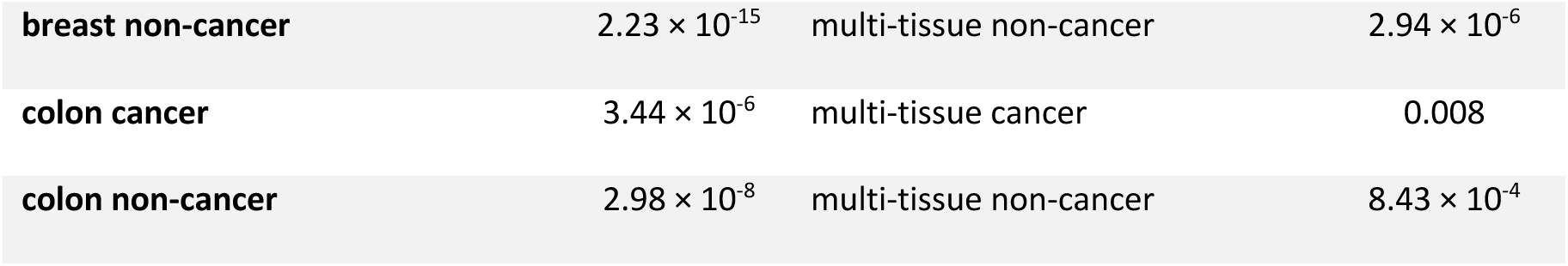
Enrichment of cancer-specific somatic driver genes for multi-tissue networks applied to cancer GWAS summary statistics.

Overall, the best performing combination was the breast cancer GWAS when using the adipose- breast co-regulation network (p-value: 2.23 × 10^-15^), while the worst performing combination was the skin cancer GWAS, for which we observed no Bonferroni-significant enrichment (p-value < 1.25 × 10^-3^, calculated by dividing 0.05 by the 40 tested networks) using any of the matching networks. We additionally performed the same analysis using sets of genes with oncogene or tumour suppressor evidence as defined by the COSMIC CGC and intOGen metadata. The enrichment observed for the breast cancer GWAS was higher for tumour suppressor genes while for the prostate cancer GWAS we observed higher enrichment for oncogenes. This suggests that both oncogenes and tumour suppressor genes may be the downstream of the risk variants captured by cancer GWAS and that different tumour types may favor different types of driver genes (Fig. S6, Fig. S7).

Furthermore, for the breast cancer GWAS, we observed Bonferroni-significant enrichments (p-value < 1.25 × 10^-3^, calculated by dividing 0.05 by the 40 tested networks) for all other non-matched networks (Figure 3). We reasoned this could be due to cancer driver genes that are shared with other cancer types and not specific to breast cancer. We therefore repeated the analysis using only breast cancer specific cancer driver genes and observed that the magnitude of non-matched enrichment was diminished, showing -log10(p-values) with a range of 3.5 to 10 for non-exclusive breast cancer driver genes compared to a range of 0.5 to 3 for exclusive breast cancer driver genes (Fig. S8). It could also be the case that the enrichment observed with non-matching networks is driven by the GWAS, independently of what network is used. We therefore compared the gene-prioritisation scores between all tested multi-tissue and tissue-specific networks. For the enrichments derived from the breast cancer GWAS and all tested co-expression networks no Pearson’s correlation above 0.4 between any combination was observed (Fig. S9). This indicates that the high enrichment in non-matching networks in breast cancer GWAS is not driven by the breast cancer GWAS. For the case of GWAS paired with non-matching networks we observed the most significant enrichment for the breast cancer GWAS with the prostate networks (cancer prostate network p-value: 3.10 × 10^-8^, non-cancer prostate network p-value: 1.03 × 10^-10^, Figure 3). This observation is in concordance with a recent study that quantified shared heritability from multiple different cancer GWAS, where a positive genetic correlation was observed between breast and prostate cancer (30). Together, these results suggest that enrichment in non-matched networks is driven by cancer driver genes that are shared between multiple cancer types or shared heritability.

**Figure 3:**
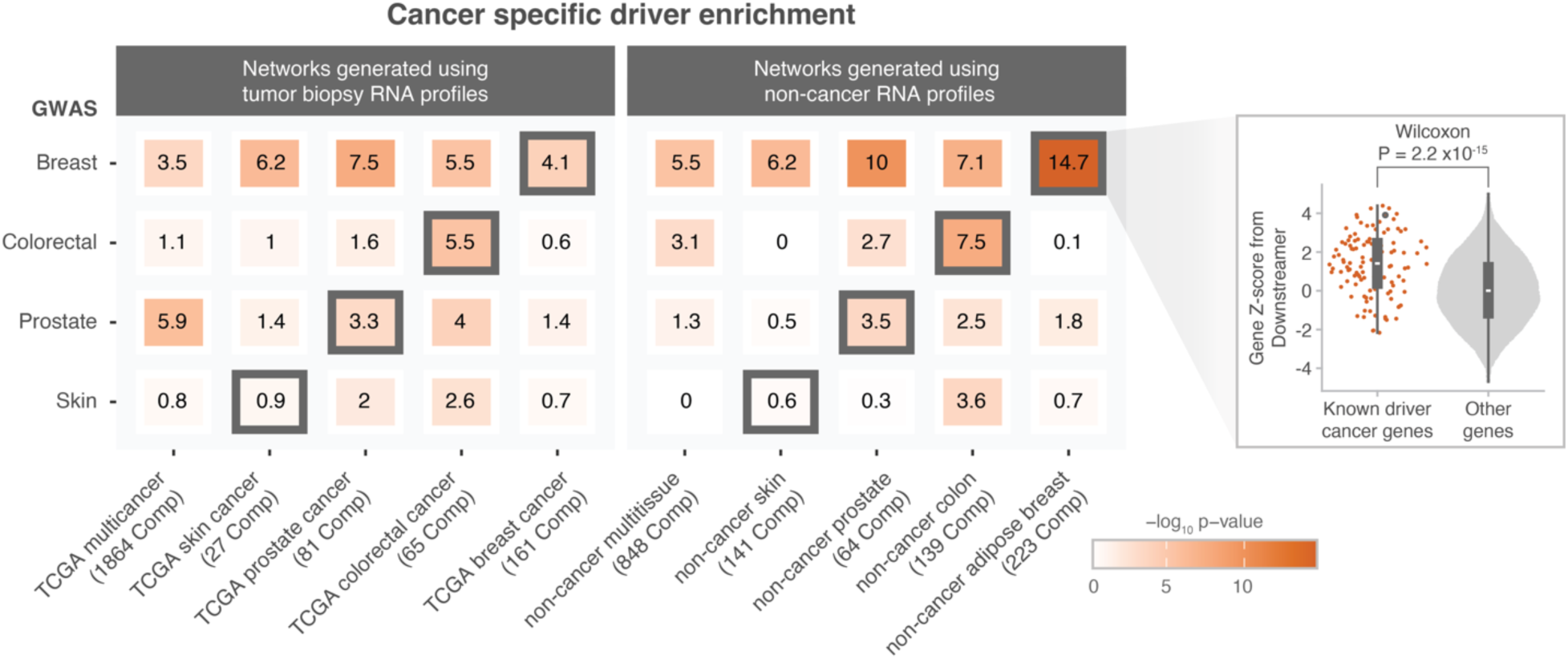
Enrichment of cancer-specific somatic driver genes for all combinations of different multi-tissue and tissue-specific gene-prioritisation methods applied to cancer GWAS summary statistics. X-axis indicates the different tissue networks created based on cancer tissue data from TCGA and the different non-cancer tissue networks created based on tissue data from recount3. Y-axis indicates the different GWAS considered in our study. A one-sided Wilcoxon signed-rank test was used to calculate the enrichment of cancer-specific somatic driver genes in the list of prioritised genes. Rectangles with a black border correspond to the tissue-specific gene-prioritisation method that matches the tissue of origin for the cancer GWAS trait. The violin chart on the right highlights an example comparison for the breast cancer GWAS results in the non-cancer adipose breast network.

### Relevant breast cancer driver genes are co-expressed with breast-cancer GWAS associated-genes through a non-cancer adipose-breast network

As shown in Figure 3, the combination of using a non-cancer adipose-breast network for the breast cancer GWAS resulted in the most significant observed enrichment in known cancer driver genes (p-value: 2.23 × 10^-15^). This network is derived from adipose tissue samples in addition to breast cancer tissue samples because these samples of breast and adipose tissue were indistinguishable during the preparation of tissue specific networks (Note S2). To showcase specific examples of genes we investigated the known cancer driver genes that were prioritised by Downstreamer to be in the top 100 ‘core’ genes.

For the non-cancer adipose-breast network that we applied to the breast cancer GWAS, Downstreamer prioritised seven known breast cancer driver genes among the top 100 genes: *CREBBP*, *TRPS1*, *ARID1A, ARID1B*, *XBP1*, *SPEN* and *NUMA1* (Figure 4). *ARID1A* and *ARID1B* are well established Tier 1 COSMIC GCC breast cancer drivers and are located far away from the index variants in the corresponding GWAS (8.2 and 4.2 Mb respectively). *TRPS1* and *XBP1* have also been statistically implicated to likely be a breast cancer driver gene based on analyses of whole tumour genome by Dietlein *et al*. (8). The *TRPS1* gene encodes a transcription factor of the GATA family that has preliminary evidence of potential driver role as a regulator of the downstream targets of the estrogen receptor *α* (31,32). *CREBBP*, *NUMA* and *SPEN* have not been linked to breast cancer by the COSMIC CGC but have been statistically determined to be breast cancer drivers by intOGen. The protein encoded by the *SPEN* gene acts as an estrogen receptor cofactor and has been shown to have a tumour-suppressor role in regulating tumour growth, cell proliferation and survival in ERα-expressing breast cancer cell lines (33). In conclusion, these results indicate that a non-cancer adipose breast network can be applied to breast cancer GWAS summary statistics to identify ‘core’ genes that are enriched for relevant somatically mutated driver genes.

**Figure 4:**
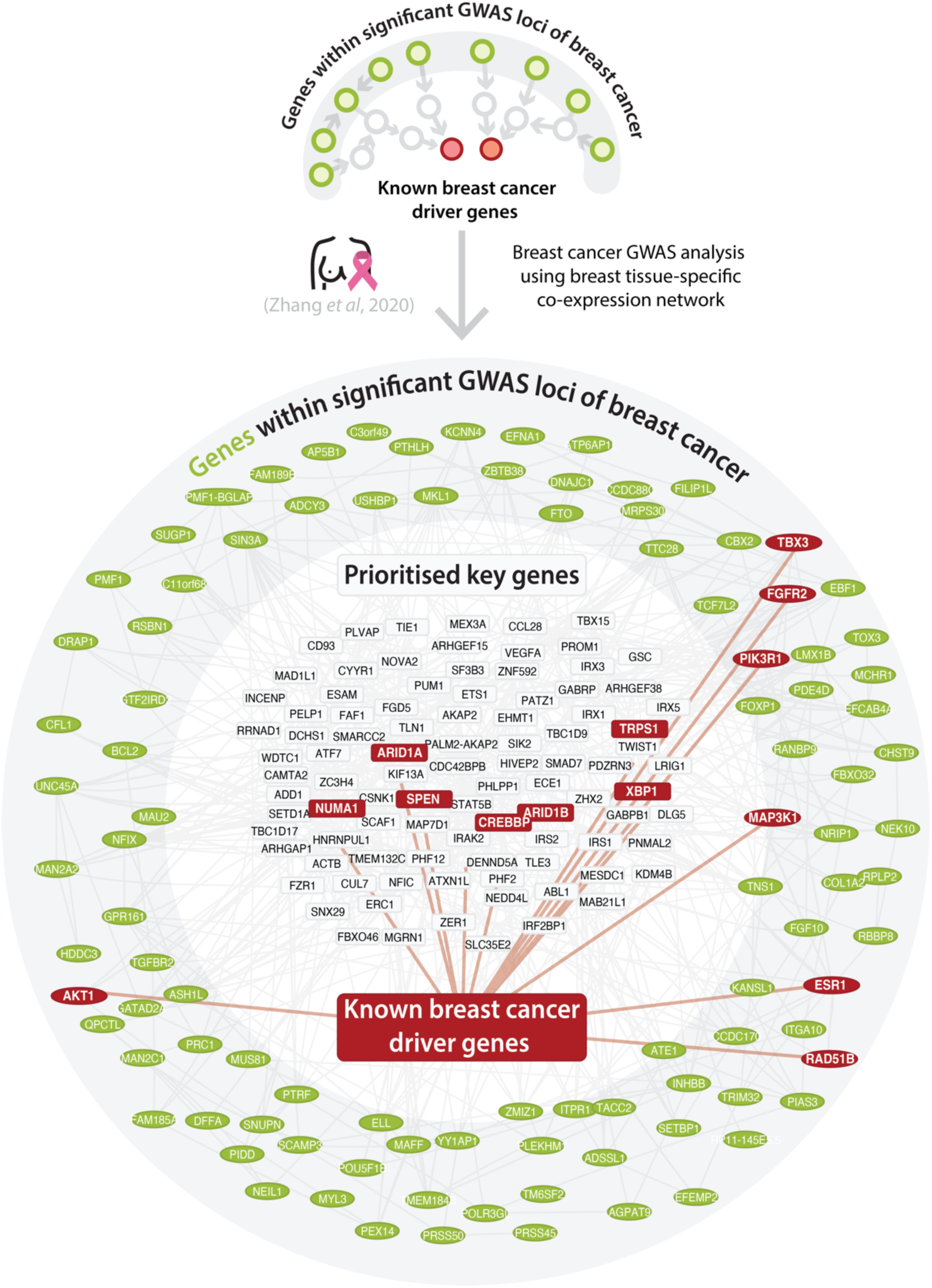
Co-expression of the top 100 prioritised genes of the breast cancer GWAS using PascalX (outer circle) and a non-cancer cancer adipose-breast tissue-specific co-regulation model with Downstreamer (inner circle). Genes in the periphery are derived from the GWAS summary statistics using PascalX (potential peripheral genes; green nodes) while genes in the centre are Derived with Downstreamer (potential ‘core’ genes; grey nodes). Edges are drawn when the co-regulation between genes has an absolute Z-score greater than two. Nodes with a red colour indicate known somatic cancer driver genes. Although some cancer drivers are prioritised by PascalX from GWAS summary statistics, the Downstreamer prioritisations point to new breast specific cancer drivers that were not prioritised using only the GWAS data alone. For illustration purposes, the top 100 prioritised genes by Downstreamer and the top 100 PascalX genes prioritised genes by Downstreamer that are Bonferroni significant in their PascalX score are shown here.

To better understand which processes are shared between the genes that are prioritised by Downstreamer we performed pathway analysis. We performed pathway analysis for the non-cancer adipose-breast network on Downstreamer-prioritised genes (5% FDR-significant) against all other tested genes. Using a Fisher’s exact test on Gene Ontology (GO) biological process terms showed a Bonferroni-significant association (p-value < 4.04 × 10^-6^, n-tests = 1,236) with Downstreamer-prioritised genes for the GO terms “regulation of transcription by RNA polymerase II” (p-value: 1.27 × 10^-7^), “positive regulation of transcription by RNA polymerase II” (p-value: 4.06 × 10^-8^) and “negative regulation of transcription by RNA polymerase II” (p-value: 2.18 × 10^-6^). Performing the same analysis against GO molecular function terms found Bonferroni-significant enrichments (p-value < 1.18 × 10^-5^, calculated by dividing 0.05 by 4243 GO molecular function terms) for “DNA-binding transcription factor activity, RNA polymerase II-specific” (p-value: 3.50 × 10^-8^) and “transcription coactivator activity” (p-value: 1.27 × 10^-6^). For GO cellular component terms, there were two Bonferroni-significant hits (p-value < 2.85 × 10^-5^, calculated by dividing 0.05 by 1757 GO cellular component terms): “chromatin” (p-value: 1.21 × 10^-6^) and “cornified envelope” (p-value: 7.33 × 10^-6^). Although these significant GO terms could be relevant for cancer, we are aware that they are not specific to cancer. All the other significant GO terms for all combinations of GWAS and networks tested can be found in the supplementary data (Data S1).

### Genes that are co-expressed with cancer GWAS associated-genes are enriched for being loss-of-function intolerant

Within the omnigenic model hypothesis we would expect ‘core’ genes as prioritised by Downstreamer to be depleted for germline missense mutations. For the prioritised genes, we compared their probability of being loss-of-function intolerant (pLI) in gnomAD to test if they are less tolerant for loss-of-function variants than other genes (34). Comparing the Downstreamer gene-prioritisation scores against the loss-of-function Z-scores provided by gnomAD using a Pearson’s correlation test highlighted a significant correlation for the colorectal cancer GWAS using the non-cancer multi-tissue network (Pearson’s *r*: 0.12, p-value: 2.31 × 10^-47^) and for the prostate cancer GWAS using the cancer multi-tissue network (Pearson’s *r*: 0.13, p-value: 2.50 × 10^-55^; Fig. S10). When using tissue-specific networks matched by the cancer’s tissue of origin, the gene-prioritisation score was significantly correlated to loss-of-function Z-scores for all combinations of cancer types and networks except the non-cancer skin network after Bonferroni-correction (p-value < 1.25 × 10^-3^, calculated by dividing 0.05 by the 40 tested networks; Fig. S11).

Overall, these results suggest that genes associated to cancer traits by GWAS may be converging on regulated cancer drivers downstream within tissue-specific regulatory networks.

## Discussion

In this study we sought to explain the limited overlap observed between genes identified through GWAS on cancer and known somatic cancer driver genes. We collected well-powered GWAS summary statistics for four cancer traits and evaluated different types of co-expression networks to ascertain if somatic cancer driver genes are potentially regulated by genes inside GWAS- cancer risk-loci. We propose that this holds for breast cancer, prostate cancer and colon cancer, particularly when using gene-networks derived from same-tissue mRNA expression data (Figure 3). These results indicate that tissue-specific gene-networks matched on the tissue of origin of the tumour can be used to link common germline variants that are associated with cancer risk to somatic driver genes. We believe this has several implications.

First, although the omnigenic model has been described in the context of germline variation between common and rare variants, our observations suggest that the model hypothesis might also apply between common germline variants and rare somatic variation. Genes inside cancer GWAS risk loci show relatively little overlap with known somatic cancer driver genes. However, we show that some cancer drivers are strongly co-regulated with genes in GWAS risk loci.

Second, our approach provides a way to use germline risk variants and gene-networks to identify potential novel cancer susceptibility genes, some of which may also have a driver gene role. This complements the strategies to search for cancer susceptibility genes, which are usually derived exclusively from cancer GWAS results and currently showing low overlap with cancer drivers (12). Future cancer subtype specific analyses using the Downstreamer methodology might confirm if cancer drivers prioritised by Downstreamer correspond to the drivers observed in that specific cancer type. This can be useful for studying the origin of the different subtypes of cancer and justifying the use of polygenic scores for patient stratification.

We also observed that cancer drivers are co-regulated with genes inside cancer-GWAS-associated loci in the context of a regulatory network derived from non-cancer tissues. This suggests that cancer driver dysregulation could occur before oncogenesis rewrites the co-expression relationships between genes. This is in line with the recent observation that non-cancer tissues harbour clonal populations of cells that have cancer driver mutations that endow these clones with a replicative fitness advantage (35).

Despite the lack of enrichment for cancer tissue derived networks, we cannot conclude that such networks are irrelevant for interpreting cancer GWAS. However, since expression of genes in cancer is highly heterogeneous, preparing cancer regulatory networks based on tissue of origin may not be ideal. Additionally, the number of cancer samples available to generate regulatory networks is lower than the corresponding set of non-cancer samples which potentially impacts power. Analyses with networks derived from more cancer samples could more confidently determine if the heritability of cancer is explained by processes that occur in the context of the altered transcription of tumours.

Our study has several limitations. Although well-powered, the GWAS of cancer traits used in this study were performed based on a clinical cancer definition that may encompass multiple oncogenesis pathways. Moreover, current estimates suggest that current cancer GWAS only explain a fraction of heritability (36), therefore this analysis can still benefit from future GWAS with bigger cohorts and with well represented cancer subtypes. Additionally, current methodologies to link associated variants within GWAS loci to genes are also not definitive and do not take tissue type into account, and future improvements of these methods could help validate links between variants and genes for different tissues. All in all, by using currently available methods, we show enrichment of cancer drivers in the Downstreamer prioritisations of core genes of some cancer-risk GWAS. In the future, with improved methodology and data, we expect that the identification of core genes for GWAS of different cancer types could lead to the identification of new cancer-susceptibility genes and cancer drivers.

While our study demonstrates the potential of co-expression networks to link cancer driver genes to cancer GWAS genes, many questions remain. For instance, we are currently uncertain if this method can be used to identify driver genes for cancers that lack identified somatic driver genes. We currently use co-expression networks derived from tumour tissue, however co-expression of genes in other cancer related cells such as immune cells could also be relevant for interpreting the cancer risk captured by GWAS.

## Conclusions

Cancer risk-variants identified through GWAS are an important resource that could serve as independent validation of the observations made when studying somatic mutation in tumour tissue. For at least some cancer tissues, common cancer risk-variants can be linked with somatic cancer driver genes using co-expression networks. This knowledge can contribute to our understanding of how cancer risk-variants contribute to the initiation and progression of cancers.

## Methods

### GWAS data

We downloaded publicly available harmonised summary statistics for 109 GWAS of cancer traits from the GWAS catalogue and from the BCAC official website (accession date: 18 October 2022) (37,38). In total, 24 different cancer traits were represented by at least one study. For each cancer, we first determined the number of independent loci in each GWAS using the *TOP_HITS* mode of Downstreamer 1.32. We then selected summary statistics with at least 90 independent loci. For every cancer trait, we selected the GWAS summary statistics derived from the study with the highest number of samples, which resulted in a final selection of four GWAS: prostate cancer, breast cancer, colorectal cancer and skin cancer (Table S1).

### Gene eigenvectors for Downstreamer

Downstreamer uses eigenvectors with gene loadings to prioritise genes. We used two sources of RNA-seq data to obtain these expression eigenvectors: recount3 and TCGA.

#### recount3 (multi-tissue)

The recount3 resource that we use to create our multi-tissue and tissue-specific networks was collected by Wilks *et al.* (21). This dataset contains 316,443 human RNA-seq samples that have been uniformly processed, covariate-corrected and quantified. We performed additional quality control (QC) and predicted if the samples are primary tissue, cell line or cancerous (Note S1).

This allowed us to select 58,725 samples expected to be non-cancer primary tissues. We then predicted the tissues of origin for samples lacking annotations (Note S2). The additional QC per tissue reduced the total number of samples to 46,410, covering 57 different tissues or cell types. We then selected the breast (n = 2,233), colon (n = 1,289), prostate (n = 386) and skin (n = 1,302) samples for the tissue-specific networks relevant to the cancers we studied.

For the multi-tissue recount3 expression matrix (n = 46,410) and the tissue-specific matrices, we used singular value decomposition on the per-gene-scaled expression data to obtain the eigenvectors with the gene loadings. For the full matrix, we selected the components that jointly explain 85% of the variance (n = 848). For the tissues, we confined ourselves to components that explain 80% of the variance and have an eigenvalue of at least 1: breast (n = 223), colon (n = 139), prostate (n = 63) and skin (n = 141).

#### TCGA

We used the TCGA (39) data as quantified by the recount3 project (21) for the multi-cancer network and the cancer-specific networks. We performed the same QC as for the recount3 data (Note S1) but did not need to predict the tissue or cancer status because the TCGA data is extensively annotated. We confined ourselves to samples that passed QC and are annotated as ‘primary tumors’ (n = 8,739). Genes expressed in <50% of the selected samples were excluded. The samples were jointly variance stabilising transformed (VST) using tumour origin as the confounder. We then applied the same covariate correction that we applied to the recount3 data (Note S2). From this jointly normalised data, we extracted the tumour-specific subsets relevant to the cancers we studied: breast (n = 1,085), colorectal (n = 392), prostate (n = 501) and skin (n = 108).

In the same manner as for recount3, we calculated the eigenvectors using an explained variance threshold of 85% for primary tumour TCGA data, resulting in (n = 1,864) components, and an 80% threshold for the tumour subsets: breast (n = 161), colorectal (n = 65), prostate (n = 81) and skin (n = 27).

### Cancer-specific drivers

Cancer drivers were obtained from the COSMIC Cancer Gene Census (7), the intOGen catalogue of gene drivers (5) and the drivers identified in Dietlein *et al.* (8). The intOGen catalogue of gene drivers (Release 2020.02.01) was obtained from the intOGen official website (37). The Dietlein *et al.* collection of coding and noncoding gene drivers was obtained from the supplementary tables of the original publication (8). The collection of cancer genes from COSMIC was obtained from the official website in May 2022 (7). We considered a gene to be a cancer-specific driver when it was called as a driver gene for that cancer type in at least one collection (Table S2). The gene symbols of these collections were then mapped to Ensembl IDs using the mapIDs formula of the R package “org.Hs.eg.db”. We then manually curated the tissue-specificity of cancer drivers by inspecting the CANCER_TYPE column for intOGen, the table title in Dietlein *et al.* and the ‘Tumor Types (Somatic)’ column for COSMIC CGN. Genes were annotated to preferentially have oncogene or tumour suppressor activity based on their COSMIC CGN (“oncogene”, “TSG”), or intOGen (“Act”, “LoF”) labels.

### Downstreamer

#### Determining linkage disequilibrium covariates

When performing a standard analysis using Downstreamer, results can be confounded by the linkage disequilibrium (LD) of genomic regions (18). To overcome this, an LD covariate was prepared using European LD scores https://github.com/bulik/ldsc (40) (Table S5). For the LD covariate, we calculated the mean LD score of the variants with a minor allele frequency above 5% that were within 25 kb upstream and 25 kb downstream of protein-coding gene transcription start sites on autosomal chromosomes. Importantly, these scores were not significantly enriched for cancer driver genes (one-sided Wilcoxon signed-rank test, p-value = 0.88), making this correction appropriate for studying cancer traits.

#### Component prioritisation

For each GWAS summary statistic and each co-regulation network, we applied Downstreamer v1.32 to prioritise the components, with the following parameters: *genepruningR*: 0.8, *referenceGenotypes*: European non-Finnish 1000 genomes samples, *variantCorrelation*: 0.95, *window*: 25000, *permutations*: 100,000, *permutationFDR*: 100, *permutationGeneCorrelations*: 10,000, *permutationsRescue*: 10,000,000, *geneCorrelationWindow: -1*, *permutationPathwayEnrichment:* 10,000, covariates: (see “Determining linkage disequilibrium covariates” section above) and the additional flags “--excludeHla –forceNormalGenePvalues -- saveExcel --regress-gene-lengths”. This resulted in a set of p-values and betas from the generalised least squares (GLS) regression model for every component.

#### Gene prioritisation

The gene-level prioritisation (*G*) was then derived by the following matrix vector product: *G = CB*, where *C* is the genes x FDR 5% significant components co-regulation matrix and *B* is the column-vector of component betas obtained from the GLS regression model. The vector *G* represents the Downstreamer prioritisations scores (Table S4).

### Pathway analysis

To better understand which diseases are associated with the top prioritised genes, we performed a pathway analysis of GO terms. We analysed all the combinations of different multi-tissue and tissue-specific gene-prioritisation methods applied to the cancer GWAS summary statistics separately. For this analysis, we focused on the 5%-FDR-significant genes with the most significant prioritisation scores from the Downstreamer analysis. We downloaded the molecular function, cellular component and biological process databases (accession date/version: 1 June 2020) and filtered out any GO term with fewer than 10 annotated genes (41,42). We determined significant GO terms by performing a two-sided Fisher’s exact test on the 5% FDR significance of Downstreamer genes versus inclusion of genes in GO terms.

### Gene constraint analysis

To identify further characteristics of the genes prioritised by Downstreamer, we performed statistical tests on the Downstreamer gene score versus the probability of a gene being loss-of-function intolerant. For this statistical test, we performed a Pearson’s correlation test on loss-of-function Z-scores provided by gnomAD (34). A positive loss-of-function Z-score suggests intolerance to loss-of-function variants. Negative loss-of-function Z-scores indicate genes that have more loss-of-function variants than expected.

## Supporting information

Data S1 Pathway analysis - Cellular Component

Data S1 Pathway analysis - Molecular Function

Data S1 Pathway analysis - Biological Process

## Data Availability

All data produced in the present work are contained in the manuscript with the exception of the generated tissue-specific networks that are available upon reasonable request to the authors.

## Declarations

### Ethics approval

Not applicable.

### Consent for publication

Not applicable.

### Availability of data and materials

All GWAS summary statistics and RNA-seq data are publicly available. For further information, please see Note S1, Note S2 and Table S1. Information about cancer driver genes can be found in Table S2. PascalX results can be found in Table S3, and Downstreamer results can be found in Table S4.

### Authors’ contributions

- Conceptualisation: C.G.U.T., T.L., F.B., P.D., L.F.
- Data curation: C.G.U.T., K.D., M.Z., P.D.
- Formal Analysis C.G.U.T., T.L., F.B., P.D.
- Funding acquisition: P.D., L.F.
- Investigation: C.G.U.T., T.L., F.B., K.D., M.Z., P.D.
- Methodology: C.G.U.T., T.L., F.B., O.B.B., A.C., P.D., L.F.
- Software: C.G.U.T., T.L. F.B., P.D.
- Supervision: C.G.U.T., P.D., L.F.
- Visualisation: C.G.U.T., T.L., P.D., L.F.
- Writing – original draft: C.G.U.T., T.L., F.B., P.D., L.F.
- Writing – review & editing: W.Z., J.R., H.J.W.

Roles as defined by: CRediT (Contributor Roles Taxonomy)

## Acknowledgments

We would like to thank Kate Mc Intyre for editorial assistance. We appreciate the UG Center for Information Technology and the UMCG Genomics Coordination Center, and their sponsors BBMRI-NL & TarGet, for the storage and computing infrastructure.

The breast cancer genome-wide association analyses for BCAC and CIMBA were supported by Cancer Research UK (PPRPGM-Nov20\100002, C1287/A10118, C1287/A16563, C1287/A10710, C12292/A20861, C12292/A11174, C1281/A12014, C5047/A8384, C5047/A15007, C5047/A10692, C8197/A16565) and the Gray Foundation, The National Institutes of Health (CA128978, X01HG007492 - the DRIVE consortium), the PERSPECTIVE project supported by the Government of Canada through Genome Canada and the Canadian Institutes of Health Research (grant GPH-129344) and the Ministère de l’Économie, Science et Innovation du Québec through Genome Québec and the PSRSIIRI-701 grant, the Québec Breast Cancer Foundation, the European Community’s Seventh Framework Programme under grant agreement n° 223175 (HEALTH-F2-2009-223175) (COGS), the European Union’s Horizon 2020 Research and Innovation Programme (634935 and 633784), the Post-Cancer GWAS initiative (U19 CA148537, CA148065 and CA148112 - the GAME-ON initiative), the Department of Defence (W81XWH-10-1-0341), the Canadian Institutes of Health Research (CIHR) for the CIHR Team in Familial Risks of Breast Cancer (CRN-87521), the Komen Foundation for the Cure, the Breast Cancer Research Foundation and the Ovarian Cancer Research Fund. All studies and funders are listed in Zhang H et al. (*Nat Genet*, 2020).

## Supplementary data

Note S1. Quality control and sample selection of recount3 data

Note S2. Tissue prediction and per tissue quality control of recount3 data

**Fig. S1.**
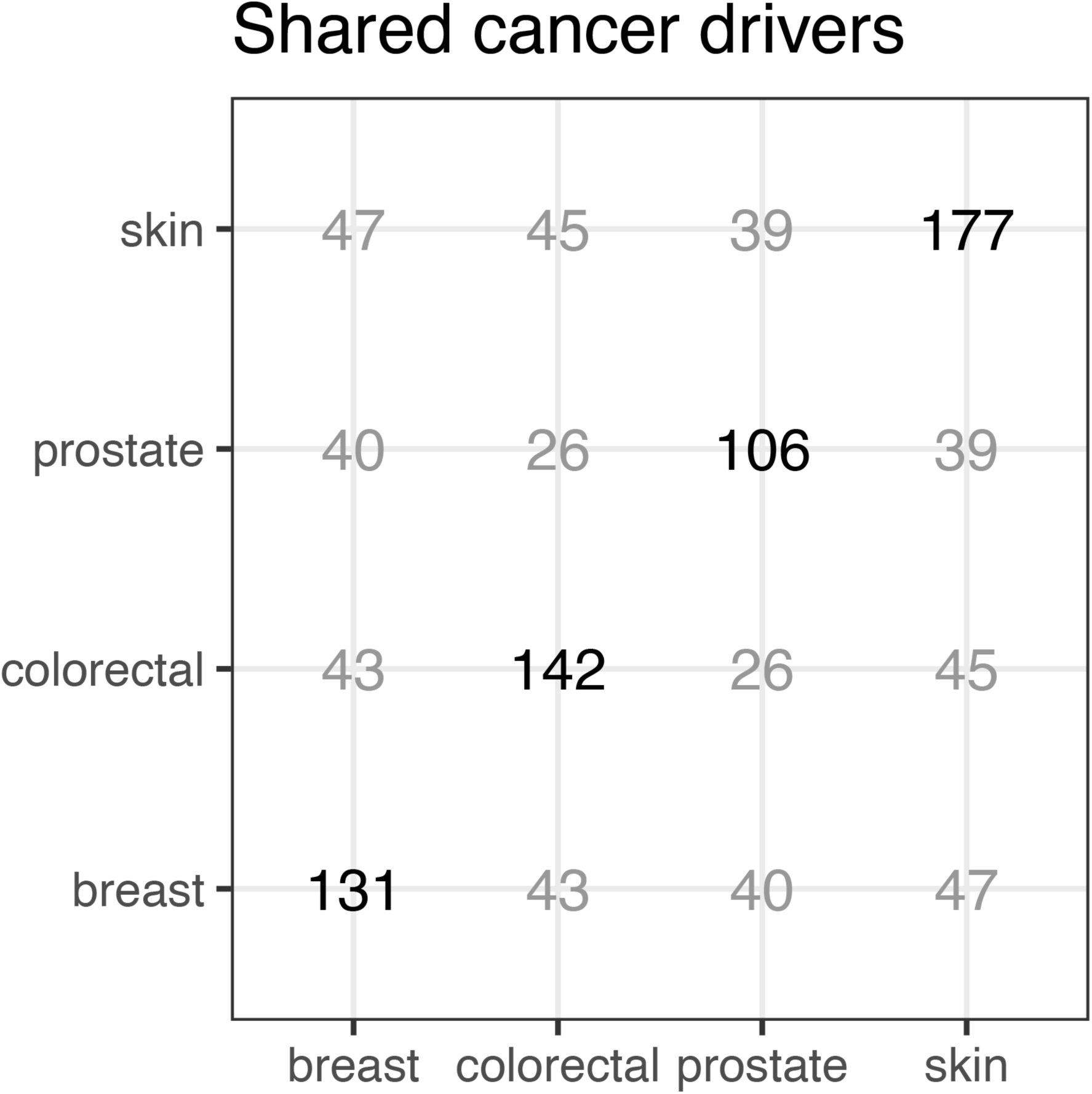
Number of cancer-specific drivers and the number shared across tumour types.

**Fig. S2.**
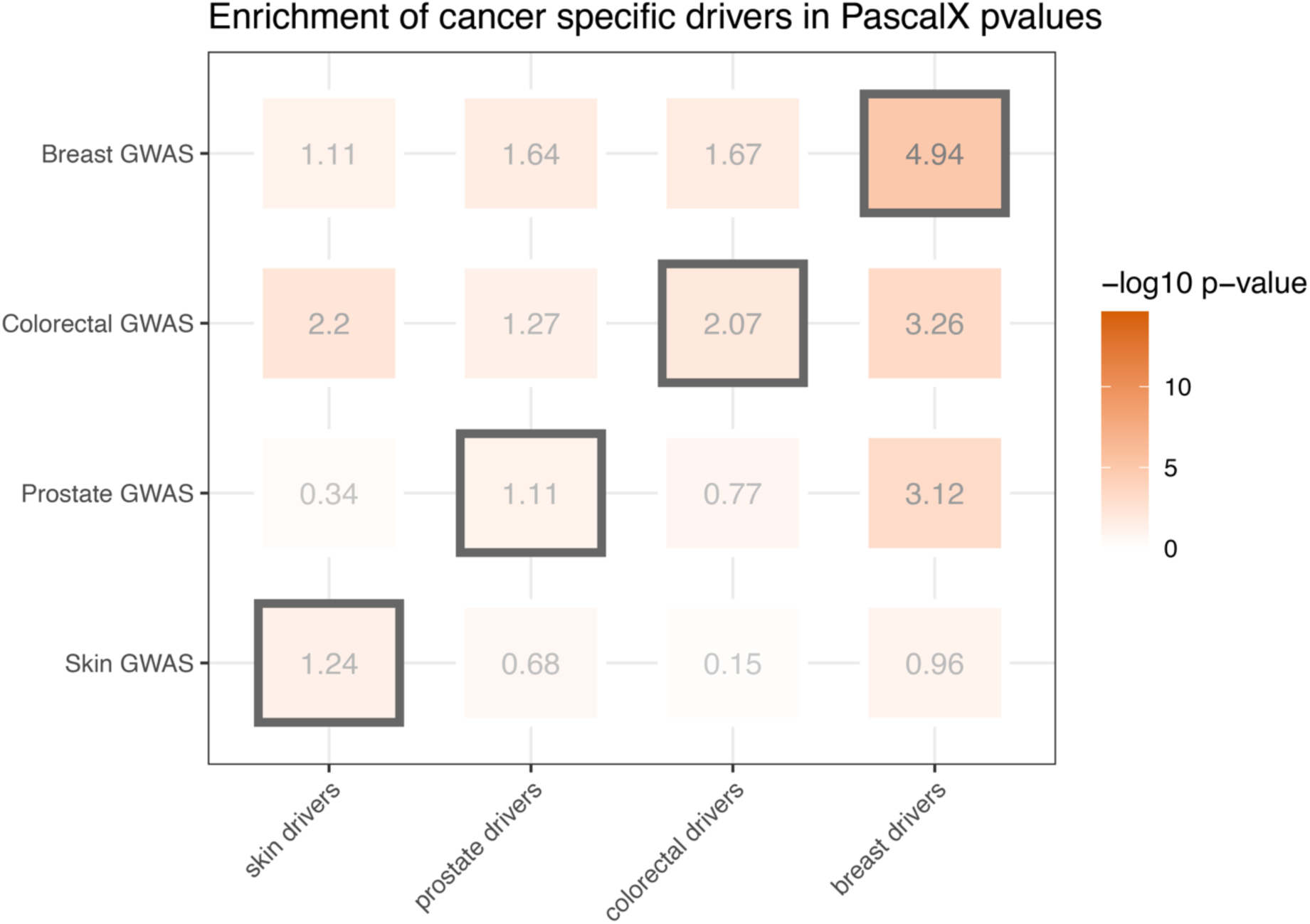
Enrichment of cancer-specific driver genes for different PascalX prioritisations.

**Fig. S3.**
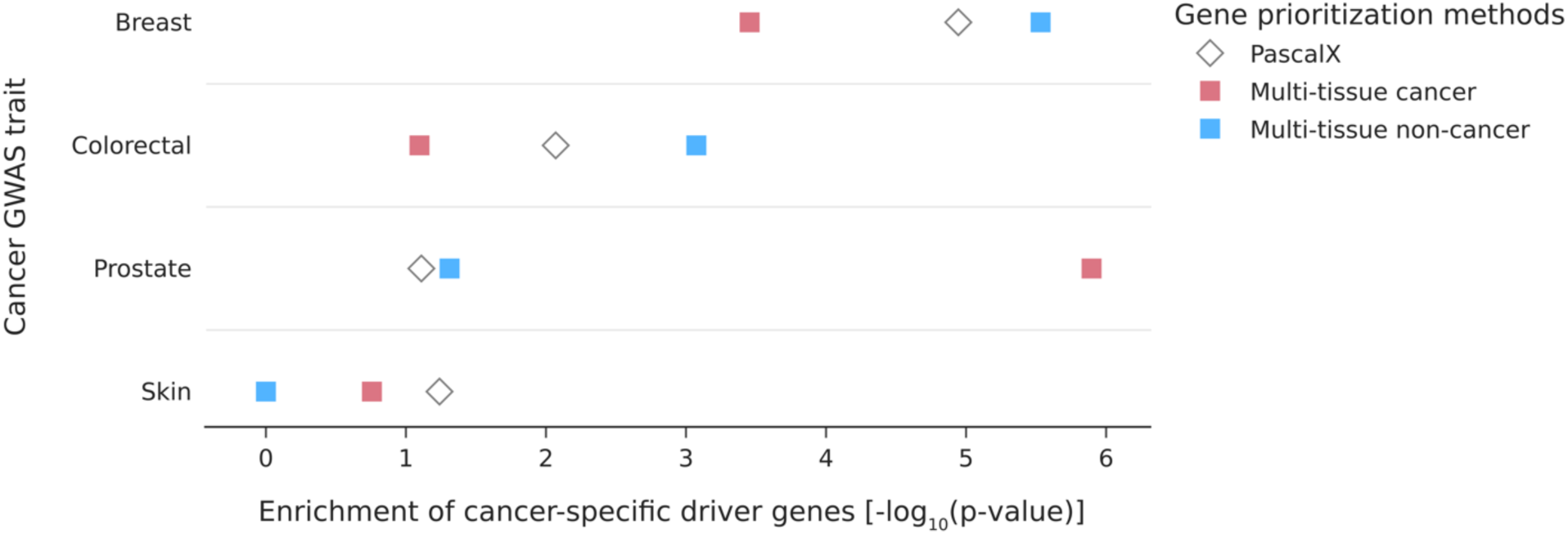
Enrichment of cancer-specific somatic and germline driver genes for different multi-tissue- and tissue-specific-based gene-prioritisation methods applied to cancer GWAS summary statistics.

**Fig. S4.**
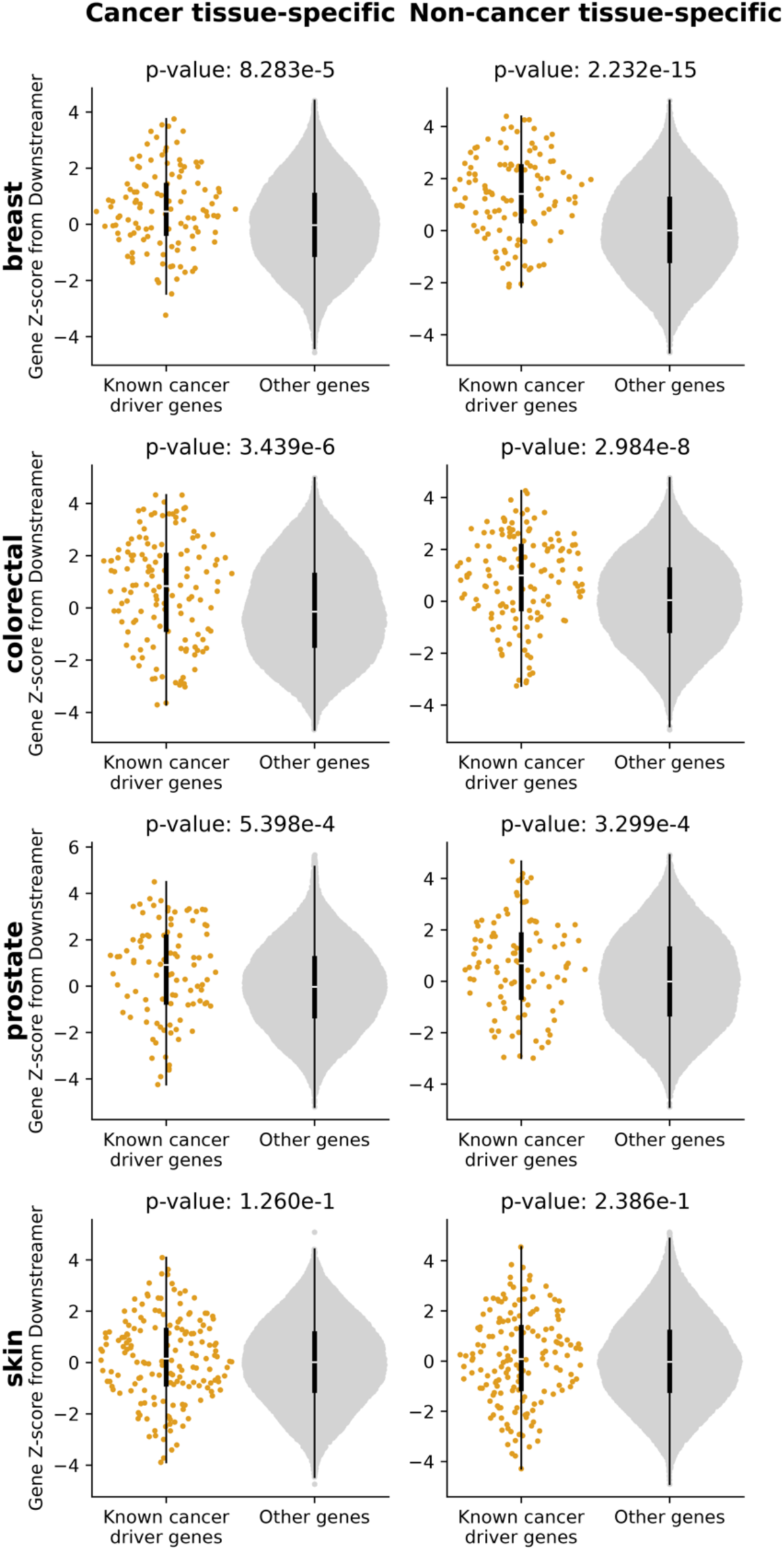
Enrichment of cancer-specific driver genes for different tissue-specific matched networks.

**Fig. S5.**
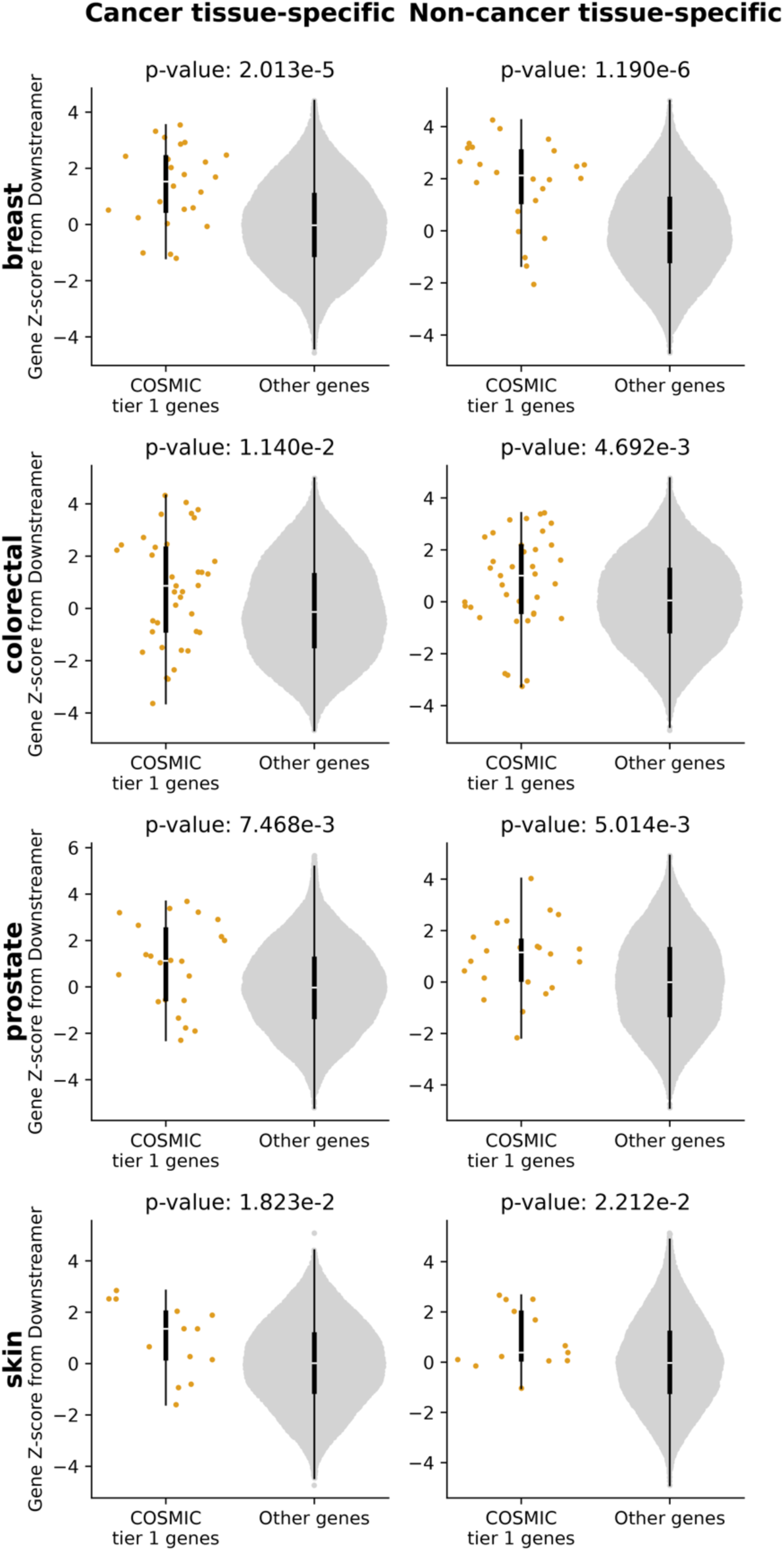
Enrichment of COSMIC tier 1 cancer-specific driver genes for different tissue- specific matched networks.

**Fig. S6.**
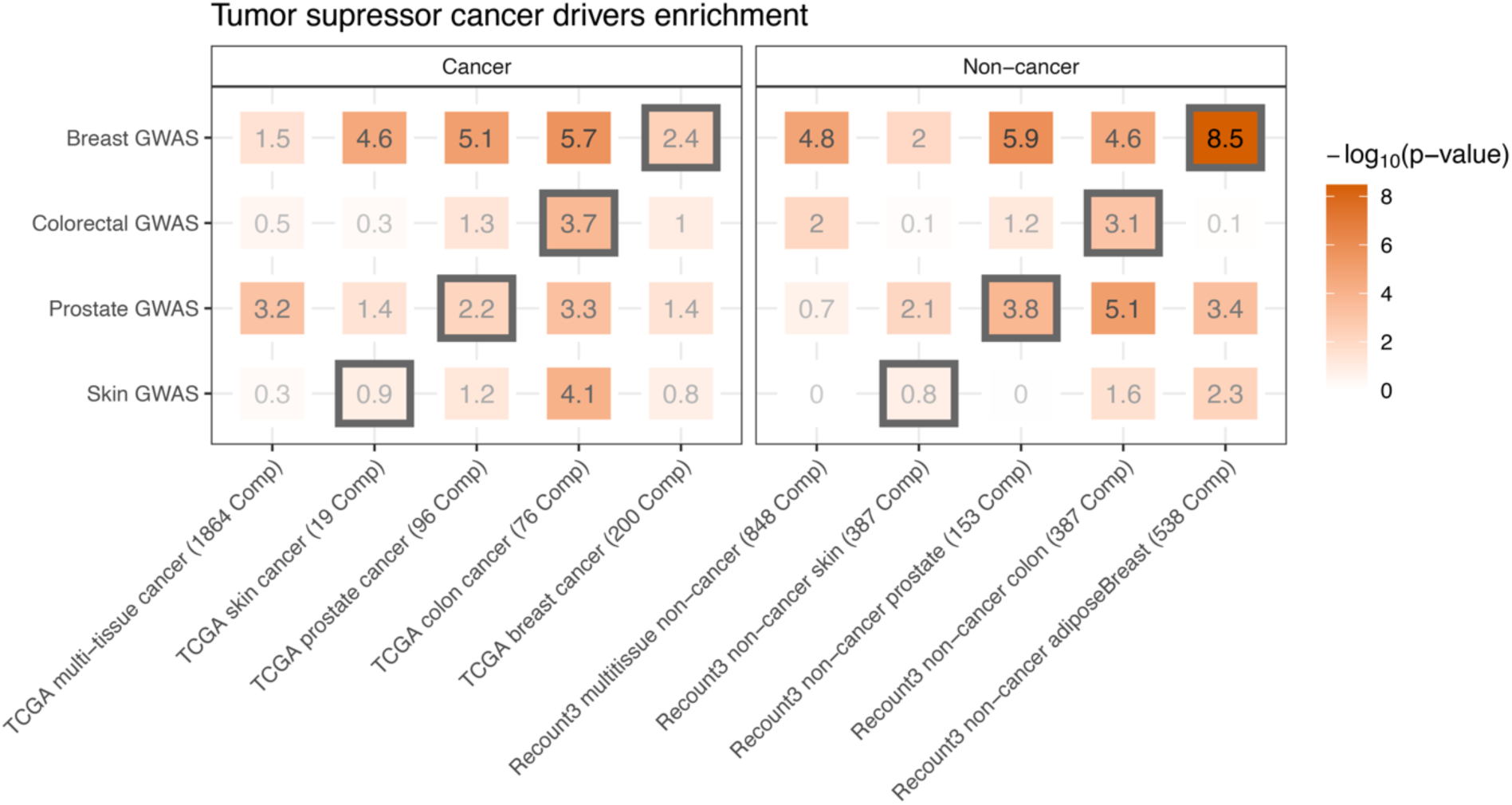
Enrichment of cancer drivers suspected to be tumor suppressors.

**Fig. S7.**
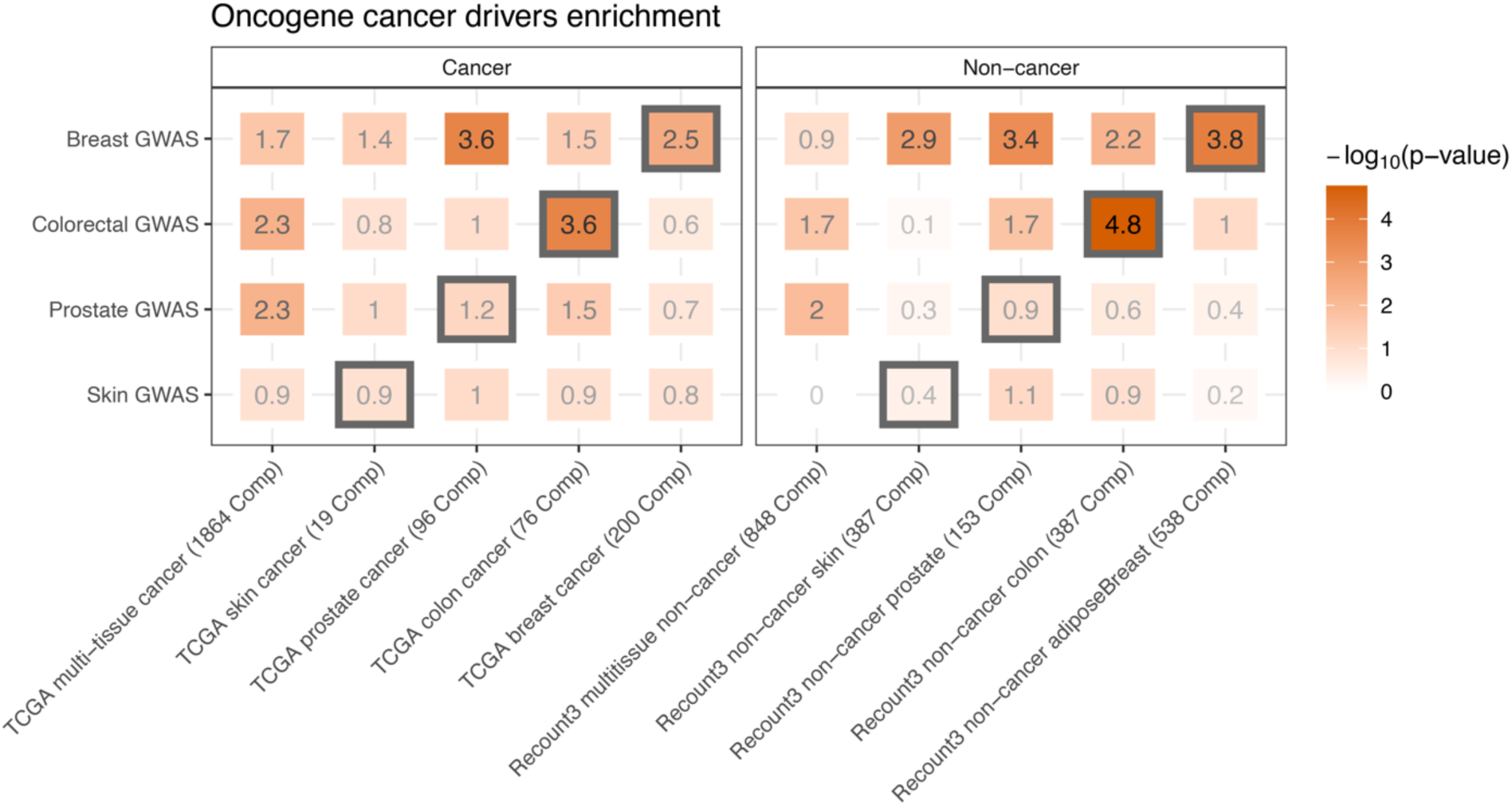
Enrichment of cancer drivers suspected to be oncogenes.

**Fig. S8.**
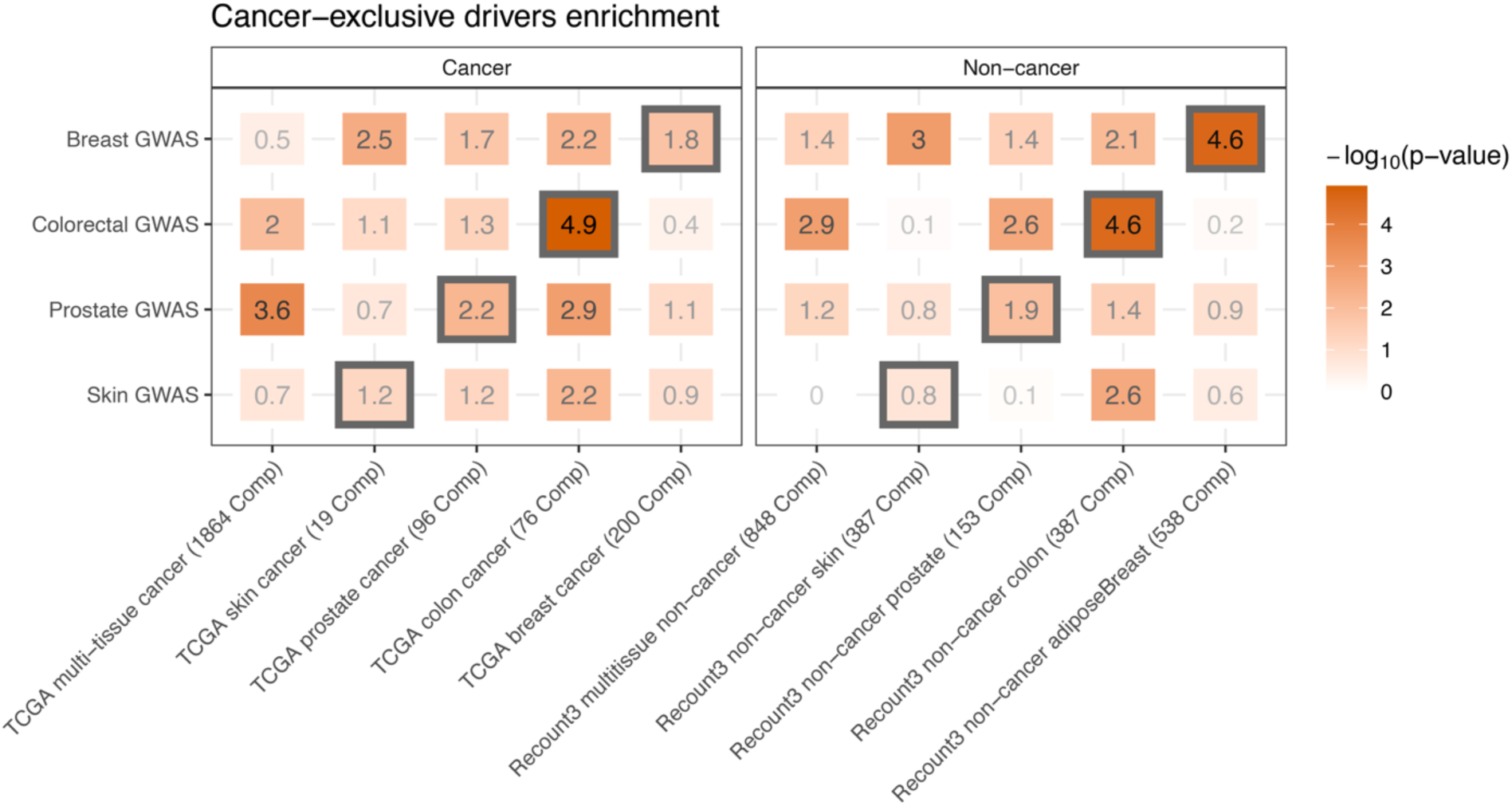
Enrichment of cancer drivers exclusive to their tissue of origin for the different prioritisation scores.

**Fig. S9.**
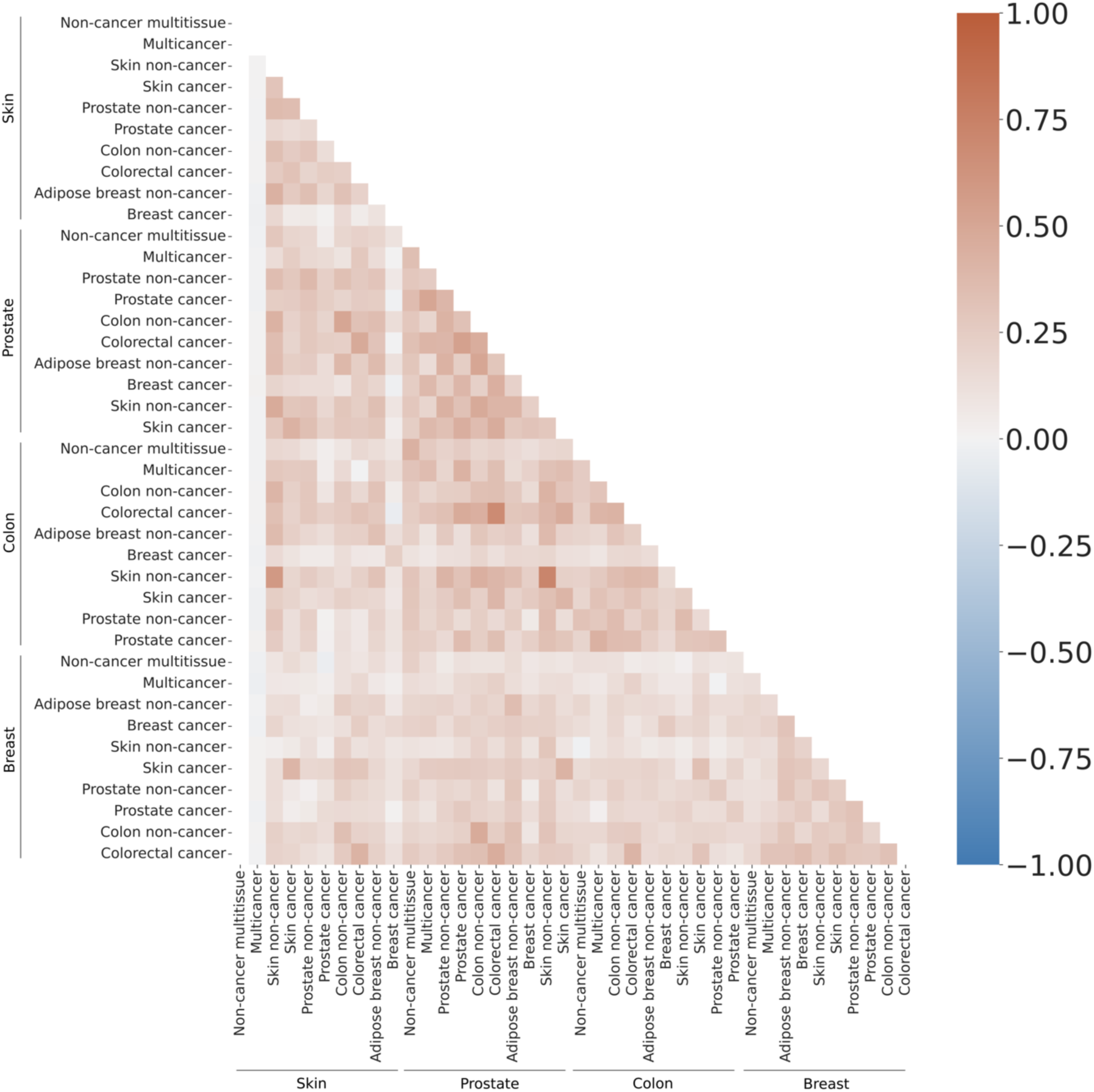
Correlations of Downstreamer Z-scores of all combinations of different multi- tissue and tissue-specific networks.

**Fig. S10.**
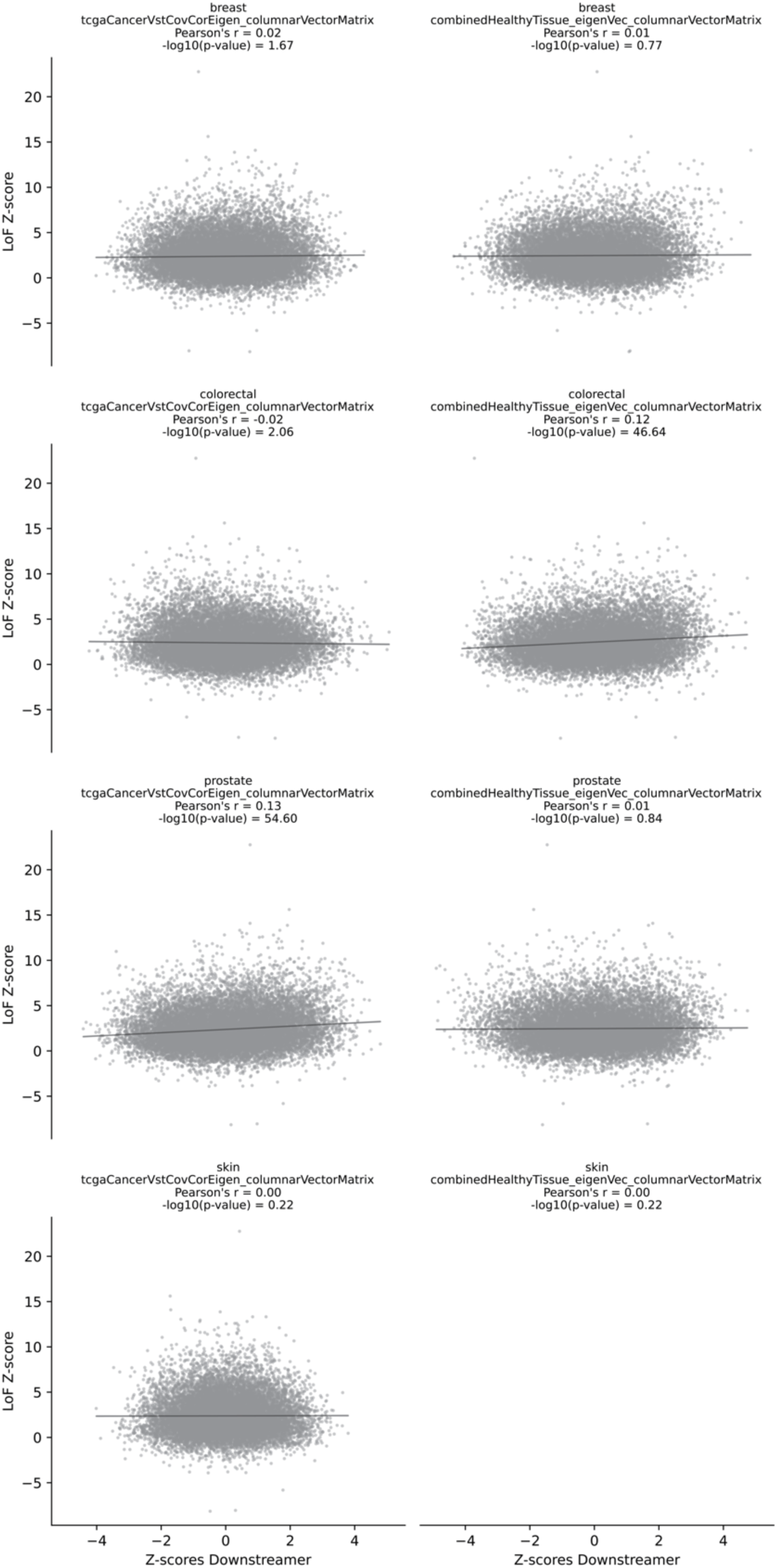
Downstreamer gene-prioritisation scores from multi-tissue networks versus loss-of-function Z-scores indicating gene constraint.

**Fig. S11.**
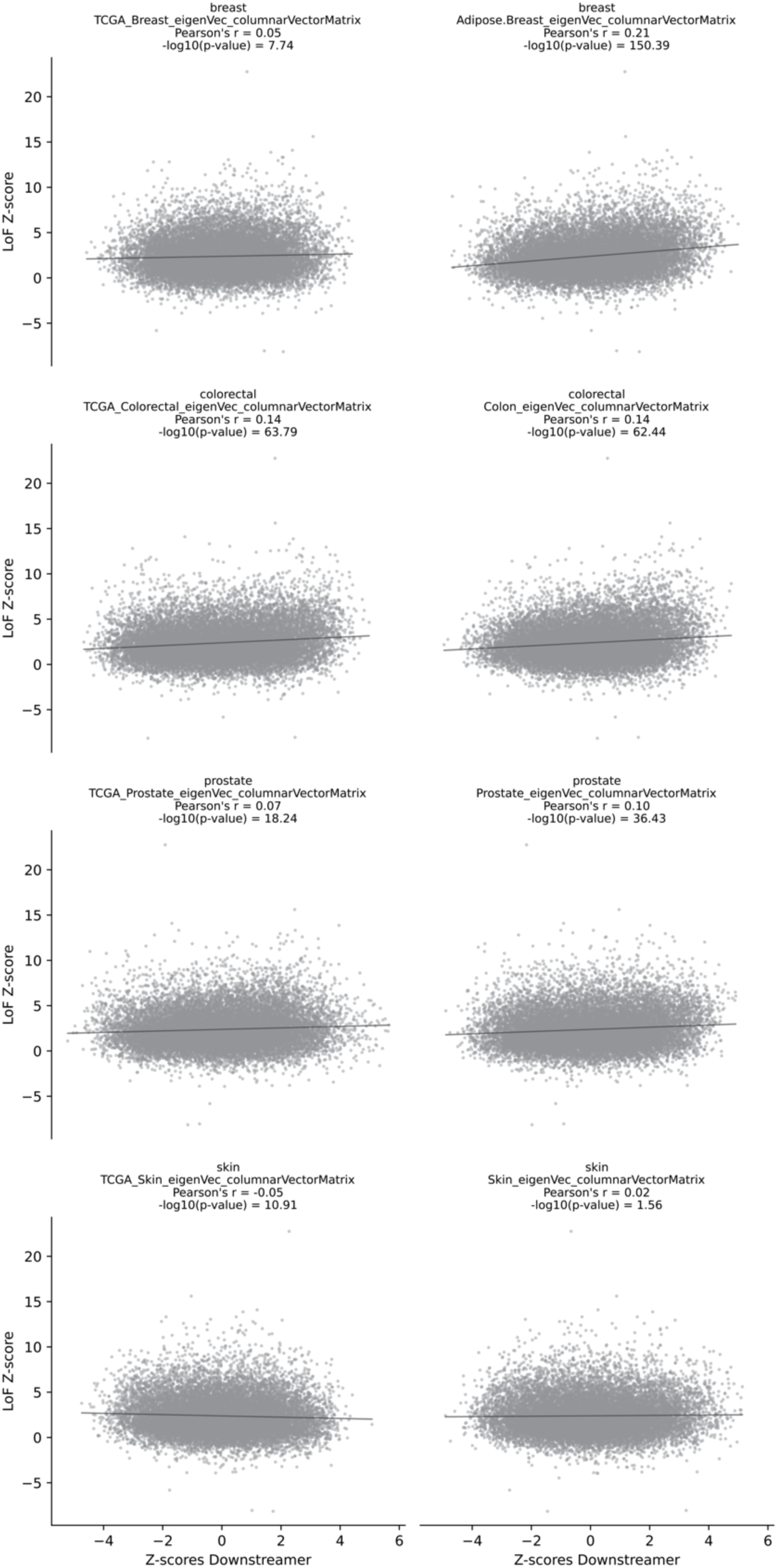
Downstreamer gene-prioritisation scores from tissue-specific networks versus loss-of-function Z-scores indicating gene constraint. *Provided separately*

Data S1. Pathway analysis

### Note S1. Quality control and sample selection of recount3 data

Phase 1: We first performed a rough selection of samples using the following steps.

- Remove samples annotated as single cell (n=74,412).
- Remove 4SU-labelled samples (n=1,589).

`o` Mention of ‘4su’ or ‘thiouridine’ in one of these columns:

“sra.library_construction_protocol”; “sra.study_abstract”; “sra.experiment_title”;
“sra.design_description”; “sra.sample_description”;
“sra.library_construction_protocol”; “sra.sample_attributes”; “sra.sample_title”
- Remove samples with only NaN expression values (n=239).
- Remove samples with missing metadata (n=1,711).
- Exclude samples based on the following quality control (QC) metrics (n=96,597):

`o` sra.sample_spots <1E6 or >2e8

▪ for TCGA samples, recount_qc.bc_frag.count <1E6 or >2e8
`o` recount_qc.star.uniquely_mapped_reads_% <60%
`o` recount_qc.aligned_reads%.chrm >20%
`o` recount_qc.aligned_reads%.chrx >6%
`o` recount_qc.aligned_reads%.chry >0.5%
`o` recount_seq_qc.%n >2%
`o` recount_seq_qc.%a <20% or >35%
`o` recount_seq_qc.%c <20% or >35%
`o` recount_seq_qc.%g <20% or >35%
`o` recount_seq_qc.%t <20% or >35%
`o` recount_qc.star.%_of_reads_mapped_to_too_many_loci >0.5%
`o` recount_qc.junction_count >500,000
`o` recount_qc.star.deletion_average_length >3
`o` recount_qc.star.number_of_splices:_total <150,000
`o` recount_qc.intron_sum_% >20
`o` recount_qc.bc_auc.unique_% <125
- Exclude all data from study SRP025982 (mixed tissues and spiked data for benchmarks).

Phase 2: We only retained genes that were expressed in at least 50% of the samples.
Phase 3: We performed another sample QC using only the maintained genes.

- Exclude samples with 0 expression >50% of the genes.
- Remove duplicate samples.
- Exclude samples with 0 variance.
- Use singular value decomposition (SVD) on quantile-normalised expression to remove outliers on the first component.

Phase 4: We corrected the remaining samples for covariates using the following steps.

- Correct the expression data for the following technical covariates:

`o` recount_seq_qc.avg_len
`o` sra.sample_spots

▪ recount_qc.bc_frag.count
`o` recount_qc.star.uniquely_mapped_reads_%
`o` sra.library_layout
`o` recount_qc.aligned_reads%.chrm
`o` recount_qc.aligned_reads%.chrx
`o` recount_qc.aligned_reads%.chry
`o` recount_seq_qc.%a
`o` recount_seq_qc.%c
`o` recount_seq_qc.%g
`o` recount_seq_qc.%t
`o` recount_qc.bc_auc.unique_%
`o` recount_qc.intron_sum_%
`o` recount_qc.star.%_of_reads_mapped_to_too_many_loci
`o` recount_qc.junction_count
`o` recount_qc.star.deletion_average_length
- 675 SRA samples were excluded for missing covariate data. Thus, the total number of samples included was 142,849.

Phase 5: We predicted cell lines and cancer samples. The predictions were based on the sample principal components and trained using the annotations known for a subset of the samples. For the prediction of primary tissues vs cell lines, we used logistic regression using the principal components.

For the prediction of cancer samples, we used the method developed by Fehrmann *et al.* (43). This first determines the auto-correlation per component, which is higher for components that reflect copy number alterations. The sample loadings are then used to create a score per sample that indicates the amount of copy number alterations in the samples. We could then use this score in a second logistic regression model that discriminated between primary tissues and cancer samples.

Neither of these models yielded perfect separation between the three classes of samples. While this is in part driven by erroneous annotations in the public repositories, further QC could improve the creation of the tissue-specific subsets.

### Note S2. Tissue prediction and per tissue quality control of recount3 data

To predict tissues for the samples that are predicted to not be cell lines or cancerous, we started anew with Transcripts per Million values. We selected the genes expressed in at least 50% of the samples, performed log2 and quantile normalisation and corrected for the same covariates as before. We then performed a new principal component analysis (PCA) and used the components in a multinominal logistic regression model trained on the known sample annotations.

One major confounder with tissue type is the associated study. Typically, samples from the same study are sequenced using the same type of sequencer and read length, and most studies investigate a single tissue. But there are many differences among the different studies. We can correct for these to some extent by including technical differences as confounders, but we found that this adversely affected our prediction accuracy. We therefore devised the following strategy to create a representative training set. Ideally, we would only use a single sample per tissue from each study to train the prediction model. In practise, for some tissues, this would result in a rather limited number of usable samples. To overcome this, we increased the number of samples per tissue per study to ensure at least 50 training samples per tissue. Based on early tests, we noticed that we could not reliably discriminate between adipose and breast samples. These samples were therefore combined in a single adipose-breast network that we refer to as a ‘breast’ network in this manuscript for clarity.

We then used the R package *glmnet* (44) to do lasso regression with cross validation to select an optimal lambda. This model was then applied to all samples, and we assigned each sample the tissue with the highest posterior probability. Samples for which the highest posterior probability was less then 0.5 were excluded.

As a final QC, we performed a PCA per tissue and excluded outliers. This resulted in 46,410 samples. Per tissue, we eventually used VST (45) for the normalisation and corrected the data for the covariates. A SVD was used to extract the eigenvectors with gene loadings that are used by Downstreamer for the gene prioritisation.

For the recount3 multi-tissue network, we used quantile normalisation and covariate correction for the 46,410 samples for which we have a predicted tissue assignment. Here we used SVD to obtain the eigenvectors.

